# Assessment of risk perception and determinants of mpox for strengthening community engagement in local populations in Cameroon

**DOI:** 10.1101/2024.03.20.24304629

**Authors:** Ernest Tambo, Pamela J. Noungoue Ngounou, Marie Paule N. Njobet, Ngo T. Tappa, Jeanne Ngogang, Mikayla Hunter, Souradet Y. Shaw, Anne W. Rimoin, Placide Mbala-Kingebeni, Jason Kindrachuk, the International Mpox Research Consortium

## Abstract

**Background:** This study assessed the current state of knowledge, including social determinants of health considerations, regarding mpox acquisition and severity in Southwest and Littoral regions, Cameroon.

**Methods:** This was a descriptive cross-sectional study carried out with mpox cases from Southwest and Littoral regions. Perceived knowledge and determinants of mpox were assessed via a self-reported questionnaire. Descriptive and inferential statistical analyses were performed.

**Results:** A total of 394 participants took part in the study. With respect to the socio-demographic characteristic of the study population, 356 (89.4%) were Cameroonians, 267 (67.1%) were females, and 261 (65.60%) were students. With regards to mpox knowledge, 278 (69.8%) of the respondents declared that mpox is caused by a virus, with 12 (3.0%) individuals responding that the signs and symptoms associated with mpox were back and/or muscle pain, skin rash, fever, pustules, and exhaustion. Knowledge scores were found to be dependent on socio-demographic background. Based on socio-behavioral determinants of mpox, 348 (87.4%) of the participants reported consumption of wild game (bushmeat) and 92 (23.1%) participants reported that mpox can be treated traditionally in their culture. Regarding epidemiological determinants, 42 (10.6%) participants reported that mpox can be transmitted through direct contact with lesions, 120 (30.2%) reported prior smallpox infection, and 47(11.8%) reported prior mpox infection. Based on risk factors reported, 180 (46.7%) of the participants had close contact with confirmed or probable cases of mpox and 196 (49.2%) were present in healthcare facilities where mpox cases were managed.

**Conclusion:** Multiple knowledge gaps regarding mpox and MPXV were documented in the population in Southwest and Littoral regions of Cameroon. Reported social and behavioral determinants included the state of instability of the southwest region and population displacement in the bushes/forests, wild game consumption without proper cooking and poor hygiene were associated with mpox infection risk perception and vulnerability. On the epidemiological aspects increased instability, travel out of town, and limited remote rural chickenpox vaccination coverage were reported to increase risk, vulnerability, and spread of mpox within these endemic communities.

## 1.0 INTRODUCTION

Mpox (formerly monkeypox) is a neglected emerging disease caused by monkeypox virus (MPXV) that can resemble smallpox presentation in humans, although associated with less severe outcomes. MPXV has emerged as the most important orthopoxvirus in public health following the successful eradication of smallpox and cessation of the global smallpox vaccination program [1, 2]. The 2022-2023 global mpox epidemic resulted in a cumulative total of 93,921 laboratory-confirmed cases, including 179 deaths, from 117 countries [3].

MPXV is an enveloped double-stranded DNA virus that belongs to genus *Orthopoxvirus*, family *Poxviridae*, and is endemic in multiple regions of Central and West Africa. Two distinct MPXV clades and two subclades have been identified: Clade I (formerly Congo Basin or Central African clade) and Clade II, which is subdivided into clade IIa (formerly West African clade) and the recently described clade IIb [4, 5]. Infection with clade I virus is associated with more severe disease as demonstrated in both humans and animal infection models. Human infections in endemic regions are often found close to tropical rainforests with rodents being the presumed reservoir for the virus. A geographical division between the two MPXV clades has been identified in Cameroon, the only country where both clades have been shown to co-circulate [6]. However, there is little information available on the historic impact of mpox in Cameroon [6, 7].

Little is understood regarding the underlying factors for the recent global expansion of clade IIb MPXV, resulting in high burdens of disease in multiple non-endemic regions, including North America and Europe. The emergence and re-emergence of new and old infectious diseases has widely been linked to poverty and vulnerability, malnutrition and poor healthcare behavior that is more prevalent in low-income regions, including sub-Saharan Africa [8-11]. Epidemiological analysis of the 2022 global mpox epidemic demonstrated a high overrepresentation of infections in males and primarily those who identified as gay, bisexual, or other men who have sex with men, as well as higher risks of infection associated with HIV infection status [12-14]. Thus, it is critical to identify the socio-behavioral and epidemiology determinants that underscored the global mpox epidemic as well those are linked to endemicity trends in Central and West Africa. Prior assessments of the spatio-temporal dynamics of mpox in the DRC have demonstrated consistent heterogeneity [15, 16]. Infection risk has been associated with physical location (e.g. proximity to forested areas), wild game contact/consumption, contact with infectious material, and length of contact (e.g. prolonged intimate contact) [15-17].

There is an urgent need for evidence and contextual lessons to inform risk communication and community engagement to build outbreak preparedness and mitigation activities within at-risk communities. Such interventions should preferentially target at-risk and marginalized populations in endemic countries, offer equal protection to citizens independent of economic status, as well as increasing investment in equitable access to diagnostics, vaccines, and therapeutics for the most vulnerable groups and populations. Given the recent global expansion of mpox, the co-circulation of clade I and II MPXV in Cameroon, the lack of consistent surveillance reporting in country, and the potential negative impact of expanded MPXV circulation on healthcare systems, there is an urgent need to increase mpox knowledge mobilization with at-risk communities and local populations. Thus, we assessed mpox risk perception and identified specific determinants of infection for strengthening community engagement and preparedness with a focus on communities in Cameroon.

## 2.0 METHODS

### 2.1 STUDY DESIGN AND DURATION

This was a descriptive cross-sectional study design carried out for a period of ten (10) months from October 2022 to August 2023.

### 2.2 STUDY REGION

The study region includes the Southwest and Littoral Regions of Cameroon. Both regions have high concentrations of tropical rainforest with populations of 1,318,000 and 2,865,795 inhabitants, respectively. The Southwest Region includes six divisions both urban and rural areas namely (Fako (Limbe), Kupe Manenguba (Bangem), Lebialem (Menji), Manyu (Mamfe), Meme (Kumba), Ndian (Mundemba)). The Littoral Region is made up of four divisions: Mungo (Nkongsamba), Nkam (Yabassi), Sanaga – Maritime (Edea) and Wouri (Douala). These areas are primarily rural with the populations primarily comprised of farmers and hunters.

### 2.3 STUDY POPULATION

The study population included patients and their contacts in the Southwest and Littoral Regions of Cameroon. This included confirmed mpox patients hospitalized in Kumba district hospital, Tiko district hospital, and Tombel district hospital in the Southwest Region, as well as the Logbaba district Hospital in Douala, Littoral Region.

### 2.4 SAMPLING TECHNIQUE

Multistage and simple random sampling was used to choose three divisions in the Southwest Region (Meme, Lebialem and Fako), and the Littoral Region at the Logbaba health District were suspected patients and contacts were enrolled and follow up for 3 to 21 days.

### 2.5 SAMPLE SIZE DETERMINATION

The sample size was calculated using the Lorentz formula which is a formula used to calculate sample size in cross-sectional studies for a total 394 participants, with the study powered to detect a prevalence of 2-5%. The sample calculation was based on Ministry of Public Health data records of infectious diseases vulnerability and prevalence in the studied areas and mpox burden estimates [18].

### 2.6 RECRUITMENT OF PARTICIPANTS WITH ACTIVE MPOX INFECTIONS

Participants were recruited at the first clinical visit following presentation to a healthcare centre for suspected mpox infection. Recruitment was facilitated through partnerships with local community-based organizations. The household contacts presumably had no signs of illness at recruitment. Those who developed symptoms ≥14 days following recruitment would then be followed as secondary contacts were followed for duration of 21 days.

### 2.7 INCLUSION AND EXCLUSION CRITERIA

Inclusion criteria were defined by participants willingness and consent to participate in the study and participants present at the time healthcare center or hospital or at the community household of the study and visit. Exclusion criteria included participants who did not consent to participate in the study or participants who did not completely fill the questionnaire.

### 2.8 DATA COLLECTION TOOL AND PRETESTING

*Questionnaire for data collection:* After explaining the objectives of the study and obtaining written consent, participants were administered the questionnaire. No identifiers were recorded on the surveys or in the databases to conserve anonymity. *Pre-testing:* Questionnaires were pretested to ensure validity and reliability. The questionnaire consisted of four parts: 1) Socio-demographics; 2) Mpox knowledge; 3) Potential mpox acquisition activities; and 4) Mpox risk perception across age and gender. Subsequent MPXV testing and reporting was performed by the national mpox surveillance and emergency response program. Trained interviewers collected data on a (written) questionnaire; each interaction lasted for at most 7-10 minutes.

### 2.9 DATA MANAGEMENT AND ANALYSIS

*Data Management:* After collection of data, questionnaires were stored and transported. Research staff examined questionnaires daily for quality checks. Data were then entered into an electronic version of the questionnaire in a password-protected computer by trained staff. *Data Analysis:* For data were extracted based on pretexted and adopted Mpox Case Report Form, and included information on demographic, epidemiological, clinical presentation, laboratory, sexual history and practices, and clinical outcome. The data was analyzed using SPSS version 25. Data were described using frequencies and percentages for continuous variables and categorical variables as appropriate. The Chi-square test was used in bivariate analyses, examining socio-demographic factors and knowledge level.

### 2.10 ETHICAL CONSIDERATIONS

An ethical clearance was provided from the Institutional Ethical Review Board (IERB), Faculty of Health Sciences, at the University of Douala (including local authorizations from Regional Delegate and Administrator at district health areas and district hospitals in Southwest and Littoral regions).

## 3.0 RESULTS

### 3.1 DESCRIPTION OF SOCIODEMOGRAPHIC CHARACTERISTICS

A total population of 398 participants were assessed within the Southwest and Littoral regions of Cameroon with 89.4% (n=356) Cameroonian participants and the remainder of foreign decent. Of the participants, the majority (57.1%, n=227) were within the ages of 15-29 years, while only 12% were >60 years. The majority of participants identified as female: (67.1%, n=267), and reported living in villages (64%, n=254). With respect to marital status, 69.3% (n=276) of participants were single. Most participants identified as students (65.6%, n=261) with few participants identifying as farmers: 7 (1.8%). Regarding education, 239 (60.1%) participants had attained a university level while 10 (2.5%) participants had no formal education. The majority of participants identified as Christian: 365 (92%) (Table 1).

**Table 1:**
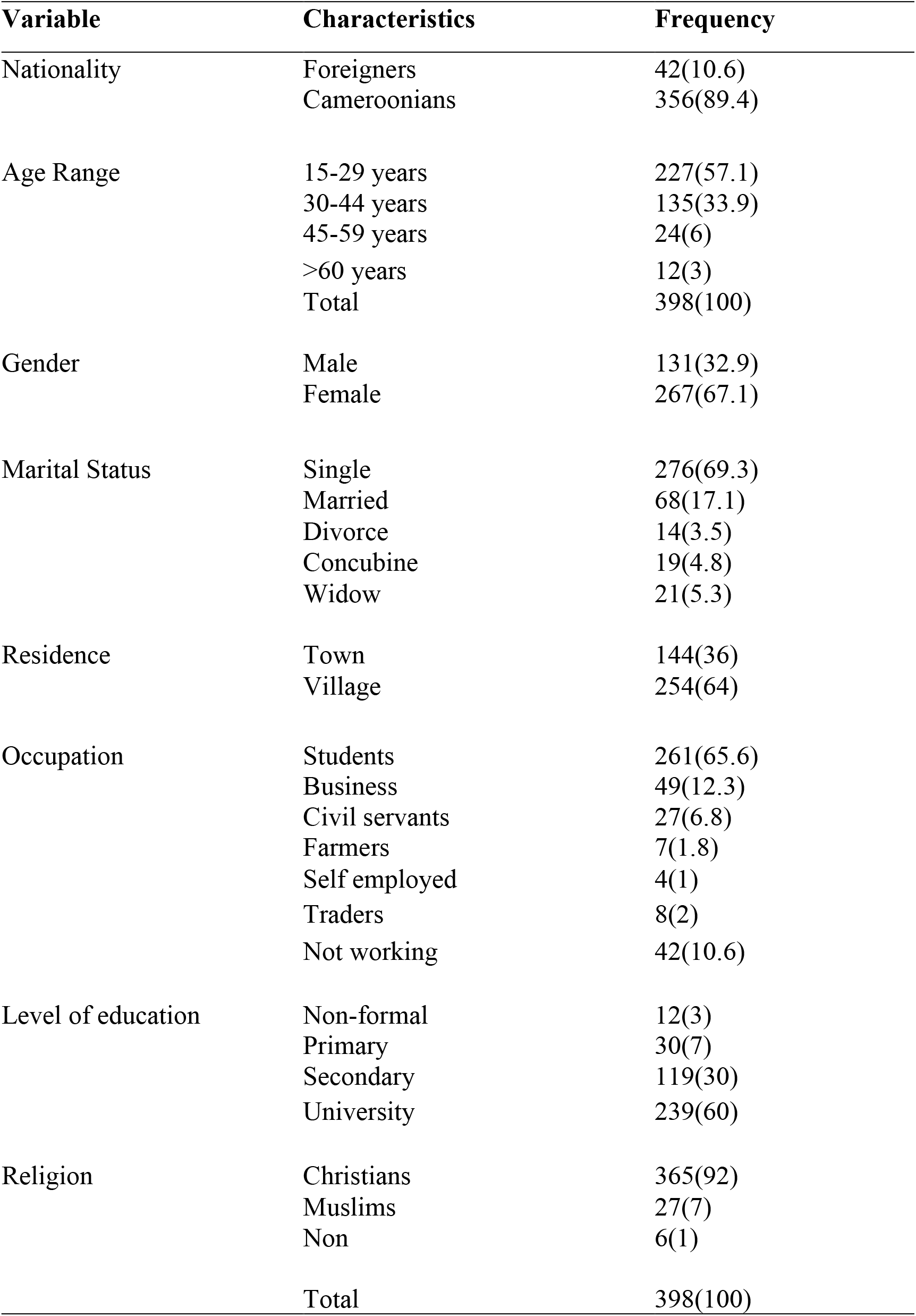
Percentage distribution of participants based on socio-demographic parameters.

#### 3.1.1 Risk perception of mpox in selected regions of Cameroon

Based on the risk factors associated with mpox, 186 (46,7%) participants reported that there was no close contact with a confirmed or probable case of mpox while the person was symptomatic. Also, 271 (68.1%) participants reported that they did not travel out of town in 14 days prior to first symptoms onset while 13 (3.3%) participants reported of not having an idea of travel out of town in 14 days prior to symptoms onset. In addition, minority participants 192 (49.2%) reported of being at a health facility where mpox was being managed. Lastly, 174 (43.7%) participants reported being present in the laboratory when handling suspected or confirmed cases as presented in Table 2.

**Table 2:**
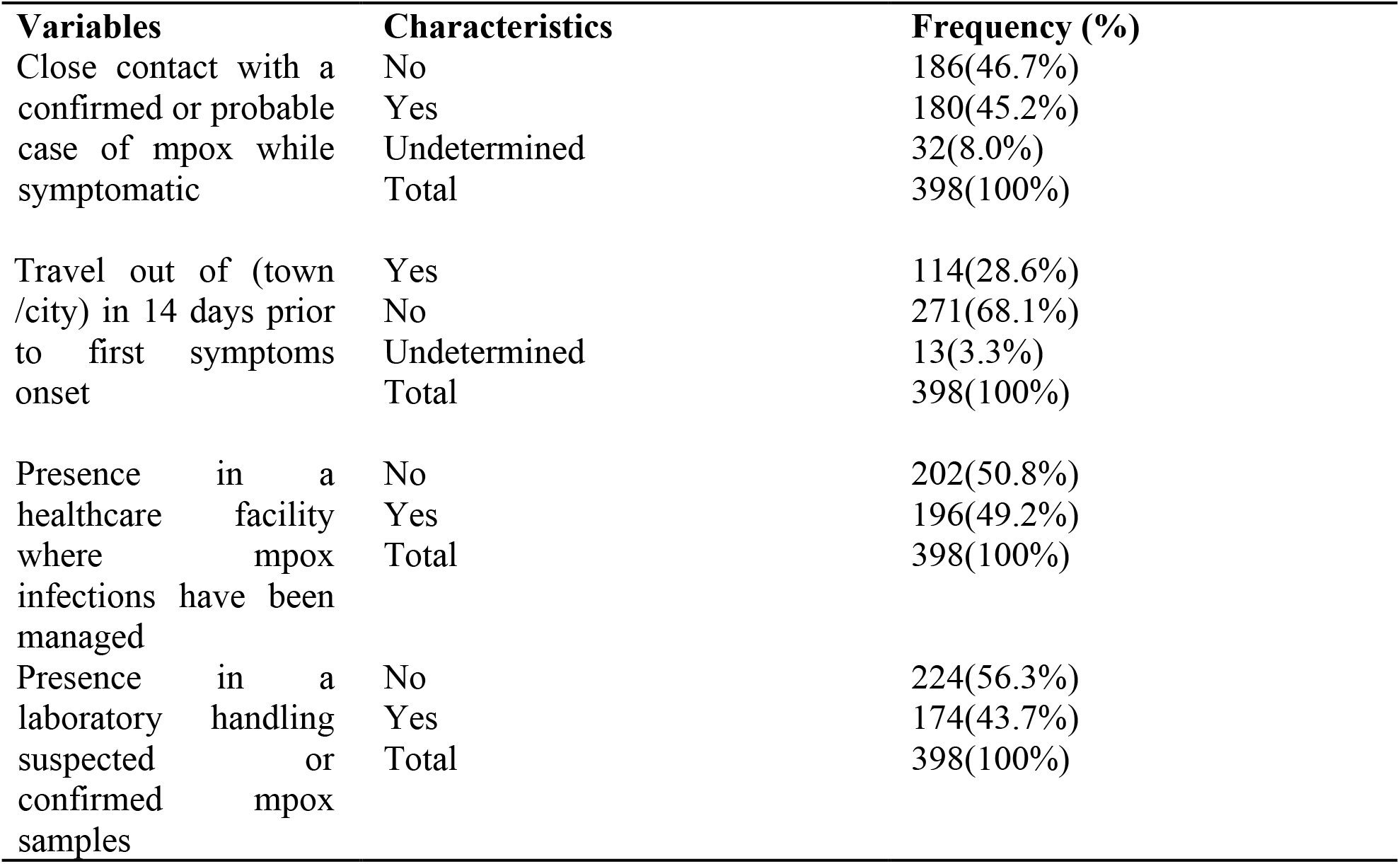
Participants distribution based on the risk factor.

| **Variables** | **Characteristics** | **Frequency (%)** |
| --- | --- | --- |
| Close contact with a confirmed or probable case of mpox while symptomatic | No | 186(46.7%) |
|  | Yes | 180(45.2%) |
|  | Undetermined | 32(8.0%) |
|  | Total | 398(100%) |
| Travel out of (town /city) in 14 days prior to first symptoms onset | Yes | 114(28.6%) |
|  | No | 271(68.1%) |
|  | Undetermined | 13(3.3%) |
|  | Total | 398(100%) |
| Presence in a healthcare facility where mpox infections have been managed | No | 202(50.8%) |
|  | Yes | 196(49.2%) |
|  | Total | 398(100%) |
| Presence in a laboratory handling suspected or confirmed mpox samples | No | 224(56.3%) |
|  | Yes | 174(43.7%) |
|  | Total | 398(100%) |

#### 3.1.2 Risk perception of viral infection

Based on the perception on the risk factors of monkeypox virus infection, 208 (52.3%) participants reported that contact with sick or dead animals was a risk of contracting mpox while 3 (0.1%) participants reported that contact with sick or dead animals, practice unsafe sex, and no PPE usage when caring for infected people (Table 3).

**Table 3:** Participants perception on risk factors of MPXV infection.

| Variables | Characteristics | Frequency (%) |
| --- | --- | --- |
| How can an individual be infected by MPXV? | Contact with sick or dead animals | 208(52.3) |
|  | Contact with sick or dead animals; practice unsafe sex; no use of PPE when caring for infected people | 3(0.1) |
|  | Contact with sick or dead animals; thoroughly cook meat; frequent hand washing | 7(1.3) |
|  | Contact with sick or dead animals; no use of PPE when caring for infected people | 14(3.5) |
|  | Contact with sick or dead animals; frequent hand washing; practice unsafe sex; no use of PPE when caring for infected people | 24(6.0) |
|  | No Idea | 15(3.8) |
|  | Practice unsafe sex | 19(4.8) |
|  | Uncooked meat | 27(6.8) |
|  | No use of PPE when caring for infected people | 42(10.6) |
|  | Infrequent handwashing | 24(6.0) |
|  | Wear face mask | 16(4.0) |
|  | **Total** | **398(100)** |

### 3.2 DETERMINANTS OF MPOX AND SEVERITY

#### 3.2.1 Social and behavioral determinants of mpox and severity

Based on the social and behavioral determinants of MPXV, out of the 398 participants 315 (79.1%) participants reported that they do not do animal farming. Also, 348 (87.4%) participants reported consuming bush meat. In addition, based on the 348 participants who consume bush meat, 110(27.6%) participants consumed bush meat monthly. In addition, majority 216 (54.3%) participants consume bush meat cooked. Moreover, 354 (88.9%) participants reported not being in contact with those infected with mpox. Still out of the 398 participants from the Southwest and Littoral recruited in this study, 196(49.2%) participants reported that fear can cause them to hesitate or deny the vaccine. Finally, 92 (23.1%) participants reported that mpox can be treated traditionally in their culture as presented in Table 4.

**Table 4:** Distribution of social and behavioral determinants of mpox.

| Variable | Characteristics | Frequency (%) |
| --- | --- | --- |
| Do you keep livestock? | No | 315 (79.1%) |
|  | Yes | 84 (21.1%) |
|  | Total | 398 (100%) |
| Do you consume wild game? | No | 50 (12.6%) |
|  | Yes | 348 (87.4%) |
|  | Total | 398 (100%) |
| If yes, how often | Weekly | 72 (18.1%) |
|  | Monthly | 110 (27.6%) |
|  | Trimesters | 65 (16.3%) |
|  | Yearly | 101 (25.6%) |
|  | Total | 348 (87.4%) |
| In which form do you consume wild game | Cooked | 216 (54.3%) |
|  | Roasted | 70 (17.6%) |
|  | Smoked | 74 (18.6%) |
|  | Roasted, cooked | 14 (3.5%) |
|  | Smoked, roasted, cooked | 24 (6.0%) |
|  | Total | 398 (100%) |
| Have you come in contact with a person infected with mpox? | No | 335 (84.2%) |
|  | Yes | 63 (15.8%) |
|  | Total | 398 (100%) |
| Are you on any family planning method? | No | 328 (82.4%) |
|  | Yes | 70 (17.6%) |
|  | Total | 398 (100%) |
| Have you received blood transfusions? | No | 341 (85.7%) |
|  | Yes | 57 (14.3%) |
|  | Total | 398 (100%) |
| What can cause you to deny the vaccine> | Fear | 196 (49.2%) |
|  | Adverse reactions | 14 (3.5%) |
|  | Poor information | 126 (31.7%) |
|  | Chronic disease | 25 (6.3%) |
|  | Poor information | 13 (3.3%) |
|  | Fear regarding adverse reactions | 14 (3.5%) |
|  | Total | 398 (100%) |
| In my culture, mpox is treated traditionally | No | 306 (76.9%) |
|  | Yes | 92 (23.1%) |
|  | Total | 398 (100%) |

#### 3.2.2 Epidemiological determinants of mpox and severity

Based on the epidemiological determinants of mpox, 139 (34.9%) participants reported that mpox cannot be transmitted through breast milk while 42 (10.6%) participants reported of the possibility of transmission through animal lesions. Also, out of the 398 participants in the study, 278 (69.8%) participants reported not previously having smallpox disease. In addition, 47 (11.8%) participants have been infected of mpox while 351 (88.2%) participants have not been infected with mpox/ Moreover, 44 (11.1%) participants have been in contact with those infected with MPXV, 13 (3.3%) participants reported that fear and poor information can also cause them to hesitate or deny the vaccine. epidemiological determinants of mpox revealed that 42 (10.6%) participants reported of the possibility of transmission through animal lesions. Also, 120 (30.2%) participants reported of having been infected of smallpox before. In addition, 47 (11.8%) participants have been infected of mpox as seen in Table 5.

**Table 5:** Percentage distribution of participants based on the epidemiological determinants of mpox outbreak acquisition and severity.

| **Variables** | **Characteristics** | **Frequency** |
| --- | --- | --- |
| Which of the following does not transmit mpox? | Animal lesions | 42(10.6%) |
|  | Animal scratches and bites | 63(15.8%) |
|  | Breast milk | 139(34.9%) |
|  | Close contact with infected person or object | 61(15.3%) |
|  | Human respiratory droplets | 58(14.6%) |
|  | No Idea | 35(8.8%) |
|  | **Total** | **398(100%)** |
| Have you been infected with smallpox before? | No | 278(69.8%) |
|  | Yes | 120(30.2%) |
|  | **Total** | **398(100%)** |
| Have you been infected with mpox before | No | 351(88.2%) |
|  | Yes | 47(11.8%) |
|  | **Total** | **398(100%)** |
| Is there an available vaccine to prevent the human mpox | No | 60(15.1%) |
|  | Yes | 286(71.9%) |
|  | No idea | 52(13.1%) |
|  | **Total** | **398(100%)** |

#### 3.2.3 Overall social determinants of mpox

Based on social determinants 348 (87.4%) participants reported that the consumption of bush meat is the most common social determinant of mpox with an overall mean score of 119 and overall percentage of 29.9%. Based on social determinants, it was demonstrated that over 70.1% of the participants do not know the social factors responsible for the acquisition of mpox. as shown in Table 6.

**Table 6:** Showing percentage distribution of participants based on the overall social determinants of mpox.

| **Variables** | **YES**<br>**N(%)** | **NO**<br>**N(%)** |
| --- | --- | --- |
| Do you rear animals? | 83(21.1) | 315(79.1) |
| Do you consume wild game? | 348(87.4) | 50(12.6) |
| Have you been in contact with infected persons? | 63(15.8) | 335(84.2) |
| Are you on any family planning method? | 70(17.6) | 328(82.4) |
| Have you received transfused blood before? | 57(14.3) | 341(85.7) |
| In my culture mpox is treated traditionally | 92(23.1) | 306(76.9) |
| **Total** | 714 | 1675 |
| **Mean** | **119(29.9)** | **279(70.1)** |

#### 3.2.4 Association between knowledge and social determinant of mpox

Based on a total of 398 participants who took part in this study, 319 participants who reported that mpox is a killer disease had not had prior contact with an infected person. The 63 participants who who reported contact with an mpox patient did not know about the morbidity and mortality associated with the disease (p-value = 0.067) indicating the lack of association between these parameters. Also, of the 63 person who have been in contact with an mpox patient, all reported that mpox can be transmitted through person-to-person contact (p-value = 0.04) indicating a significant relationship between contact and disease transmission. Further, of the 63 participants who reported to have been in contact with infected persons, 30 participants reported that mpox was preventable while 33 participants reported that mpox was not a preventable disease with no significant association at p-value of 0.05 (Table 7).

**Table 7:**
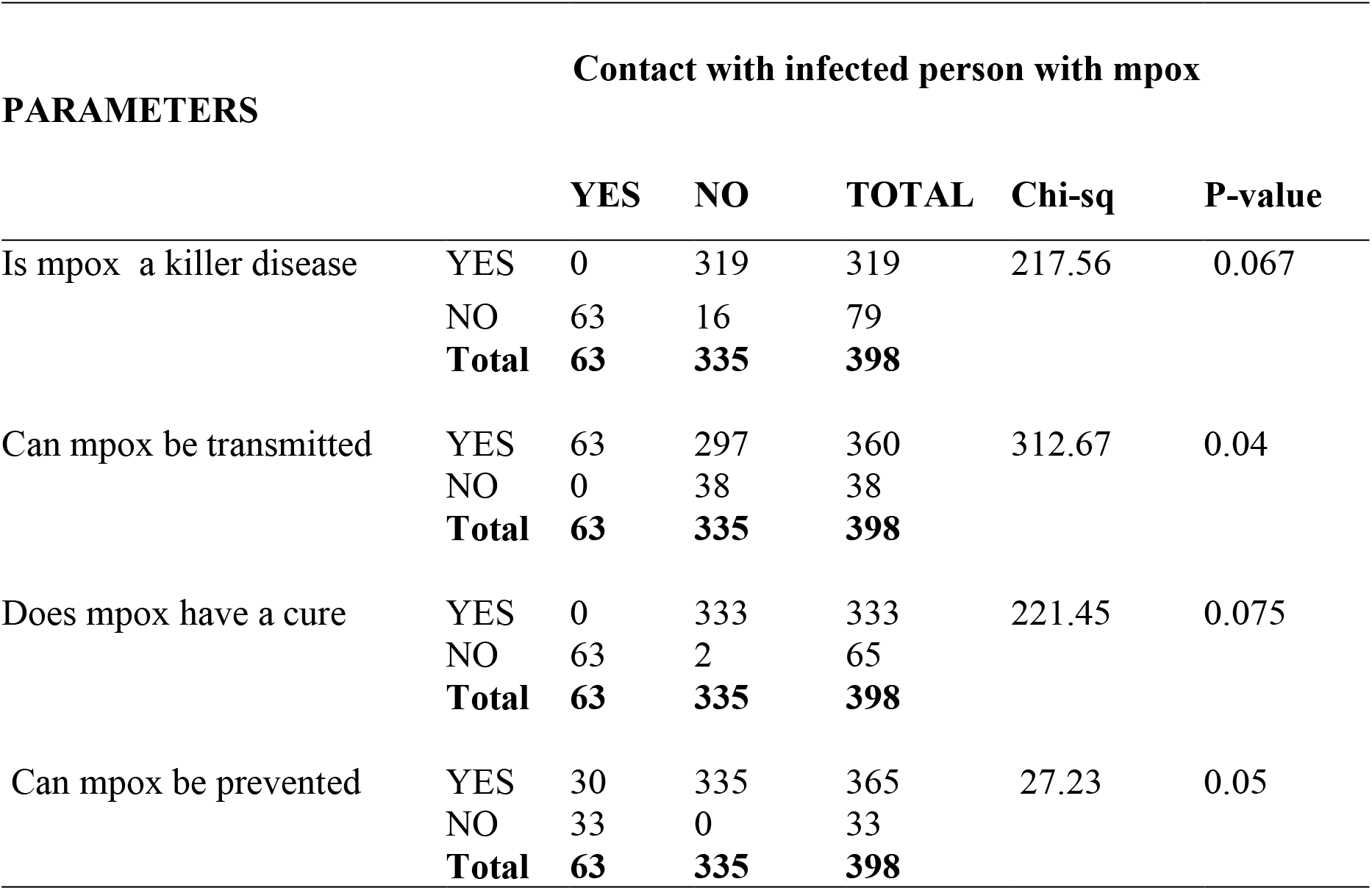
Association between Knowledge and contact with Mpox as social determinants.

## 4.0 DISCUSSION

Based on the sociodemographic characteristic, of the total participants, 131 (32.9%) were male, 227 (57.1%) fall in the age group 15-29 years, 276 (69.3%) were single, 144 (36%) of the participants residents in Urban areas and 239(60%) were at the university level. This contrasts to a study carried out by Hasham *et al* where findings revealed that of the total participants, 61.4% (n=639) were male, 61.8% (n=643) fall in age group 21–30years, 79.5% (n=827) were single. 79.5% (n=789) of the respondent residents of urban areas, and 57.2% (n= 595) achieved graduation level education [19]. This provides additional insights on the need for targeted public health and community engagement practices that are geographically specific.

Based on the risk factors associated with mpox, 186(46,7%) participants reported that there was no close contact with a confirmed or probable case of mpox while the person was symptomatic. Also, 271 (68.1%) participants reported that they did not travel out of town in 14 days prior to first symptoms onset. Additionally, 202 (50.8%) participants reported not being present in a health facility where mpox was being managed. These findings are in line with a finding which demonstrated that mpox cases involving spread among human beings are more probable among individuals who are non-vaccinated against smallpox, living in the same house, or providing care to a primary case [12]<u>[13].</u> Infected persons must be supervised to stop the further spread of the virus to vulnerable populations. Previously, WHO and others have reported on the impacts of poverty on public health and infectious diseases, including mpox [8, 10, 19, 20]. Analyzing and understanding aspects of what protections are required and how they affect individuals and societies due to under investment in health system preparedness, lack of effective mpox vaccine and therapies, weaker health systems, limited research and response capacities, and how impoverishment reflects the difficult choices facing resource-limited African countries at greatest risk. Addressing these concern would decrease the risk for undetected emergence events and/or cryptic transmission and will have global benefits in addressing early management and control of outbreaks [1, 2, 10]. Among large populations, higher fraction of disadvantages communities and increased durations of time for the introduction of interventions are found to progressively worsen outcomes, including stigmatization and social considerations [21, 22].

Regarding the knowledge on mpox risk factors, 208 (52.3%) participants indicates that mpox can be contrasted by contact with sick or dead animals with only 3 (0.1%) participants reporting the combination of two or more risk factors approaches such as contact with sick or dead animals, practice unsafe sex, non-usage of PPE when caring for infected people <u>[7,8].</u> Also, our data had similarities to previous studies suggesting that mpox circulation was likely associated with multiple and unprotected sexual activities amongst young age/adults, contact with infected patient, infected tools and products or sexually transmitted infection status [2, 7, 11, 13, 23]. The disease can be transmitted directly or indirectly through contact with infected skin lesions of contaminated patients, share towels and bedding, close sexual contact, and unprotected sexual intercourse .

Based on the social and behavioral determinants of MPXV, out of the 398 participants 315 (79.1%) participants reported that they do not do animal farming. Also, 348 (87.4%) participants reported of consuming wild game (bush meat). That lack of immunization against smallpox makes the individual more vulnerable to be infected with mpox argues for vaccine equity considerations and deployment strategies in endemic regions and resource-limited settings [1, 2, 16, 24]. These findings are similar to prior reports on the social determinants influencing global mpox circulation including global travel and trade, job loss and COVID-19 pandemic economic and social conditions effects [25]. It has long been recognized that poverty is one of the major social and economic determinants of health, hunger, ill-health and poor environment, inadequate sanitation and poor potable drinking water related vulnerability amongst at risk groups. Since the poor populations face a higher spark risk and spread related health and economic shocks, understanding why and how the poor or rich are more vulnerable to neglected tropical disease outbreak and pandemic burden is crucial.

Strengthening health system preparedness capacity is crucial in increasing access to and uptake of safe and effective mpox vaccination and therapeutic strategies against the threat of increasing mpox geographic expansion and newly reported sexual transmission of clade I MPXV [23, 24], in particular within areas at increased risk for social and political unrest.

## 5.0 CONCLUSION

In conclusion, a large majority of the population of the Southwest and Littoral regions have limited knowledge on several aspects regarding both mpox and MPXV. Regarding the epidemiological aspects associated with mpox, increase travel out of town and length of stay, mpox vaccination status or chickenpox vaccine were noted to increase the spread of mpox in the community. Lastly, the presence of pandemics such COVID-19, poverty, and lack of surveillance capacity increases the risk for mpox circulation within communities. In the southwest region of Cameroon, the geopolitical instability of this region makes it difficult for community health personnel to have access to the community to better educate this population on mpox and how it can be prevented. The situation has also caused resulted in internal displacement including to forested areas with increased exposure to wildlife, stronger reliance on wild game as a food source, and decreased capacity for cleaning and hygiene considerations, increasing potential mpox spillover and infection risks.

## Data Availability

All data produced in the present study are available upon reasonable request to the authors.

## 6.0 CONTRIBUTORS

ET, PN, NTT, NYJ, and MPN participated in the implementation and the fieldwork. PM and MPN participated in laboratory samples processing. ET, NTT, and MP participated in data assessment and reporting. ET, SYS, AWR, PM and JK supported the conception, design, and coordination of activities.

## 7.0 DECLARATION OF INTERESTS

The authors declare no competing interests.

## 8.0 DATA AVAILABILITY

Due to the sensitive nature of our survey and the involvement of human participants we did not receive approval to collect raw participant identified data from The University of Manitoba Research Ethics Board or the Cameroon Ethics and Scientific Review Committee. All survey data was anonymized at collection with no identifiable information.

## 9.0 ACKNOWLEDGEMENTS

We are grateful to all hospitals and health center authorities and participants for their cooperation. This work was supported by the International Mpox Research Consortium (IMReC) through funding from the Canadian Institutes of Health Research and the International Development Research Centre (Grant No. 202209MRR-489062-MPX-CDAA-168421) as well as a Tier 2 Canada Research Chair in the Molecular Pathogenesis of Emerging and Re-Emerging Viruses (JK) and a Tier 2 Canada Research Chair in Program Science and Global Public Health (SYS),

## Notes

### Competing Interest Statement

The authors have declared no competing interest.

### Author Declarations

An ethical clearance was provided from the Institutional Ethical Review Board (IERB), Faculty of Health Sciences, at the University of Douala, Cameroon (including local authorizations from Regional Delegate and Administrator at district health areas and district hospitals in Southwest and Littoral regions).

